# No Additional Benefit of 185 Hz versus 130 Hz at Equivalent Energy in Deep Brain Stimulation for Tremor - A randomized prospective clinical trial

**DOI:** 10.64898/2026.05.31.26354199

**Authors:** Christina van der Linden, Peer Trapp, Till A. Dembek, Charlotte Schedlich-Teufer, Gregor A. Brandt, Hannah Jergas, Gereon R. Fink, Veerle Visser-Vandewalle, Michael T. Barbe, Jan Niklas Petry-Schmelzer

**Affiliations:** University of Cologne, Faculty of Medicine and University Hospital Cologne, Department of Neurology, Cologne, Germany; Charité – Universitätsmedizin Berlin, corporate member of Freie Universität Berlin and Humboldt Universität zu Berlin, Department of Neurology with Experimental Neurology, Berlin, Germany; University of Cologne, Faculty of Medicine and University Hospital Cologne, Department of Stereotactic and Functional Neurosurgery, Cologne, Germany; Cognitive Neuroscience, Institute of Neuroscience and Medicine (INM3) – Research Center Jülich, Jülich, Germany

## Abstract

**Background:** Deep brain stimulation (DBS) of the ventral intermediate nucleus and posterior subthalamic area (VIM/PSA) in Essential Tremor (ET) and the subthalamic nucleus (STN) in Parkinson’s disease (PD) are established treatment for tremor. To achieve maximum tremor control, increasing stimulation frequency beyond 130 Hz is part of clinical practice, but lacks scientific evidence.

**Objective:** To compare tremor suppression under total electrical energy delivered (TEED)-equivalent stimulation at 130 Hz versus 185 Hz in STN-DBS for PD and VIM/PSA-DBS for ET.

**Methods:** In this prospective, double-blind study, acute DBS effects were assessed in 18 people with ET (n = 29 hemispheres), and 25 people with PD (n = 30 hemispheres). Tremor-suppressive effects, evaluated by accelerometry, were compared with TEED-equivalent stimulation at 130 Hz and 185 Hz using linear mixed-effects models, explorative pairwise comparisons, and equivalency testing.

**Results:** Linear mixed-effects models revealed no significant effect of stimulation frequency on tremor improvement in both cohorts. Pairwise comparisons showed no consistent differences in total tremor improvement with TEED-equivalent 130 Hz vs 185 Hz DBS. Post-hoc equivalence testing confirmed equivalence of stimulation frequencies under TEED-equivalent conditions within a ± 20% margin of relative tremor improvement.

**Conclusion:** This study provides Level II evidence that a higher stimulation frequency of 185 Hz does not offer additional benefit in deep brain stimulation for tremor and supports 130 Hz as the standard stimulation frequency for tremor suppression in ET and PD.

## Introduction

Tremor is a key clinical feature of Essential Tremor (ET) and Parkinson’s disease (PD) that can be effectively alleviated by deep brain stimulation (DBS) targeting the ventral intermediate nucleus and posterior subthalamic area (VIM/PSA) in ET and i.a. the subthalamic nucleus (STN) in PD. Despite remarkable advances in optimizing stimulation location and amplitude by using imaging and sensing technologies, stimulation frequency is usually set at a standardized 130 Hz.(1,2) If sufficient tremor control cannot be achieved by optimizing contact choice and titrating amplitude, clinical practice involves increasing stimulation frequency up to 185 Hz or higher.(3–6) However, this approach is largely based on expert consensus rather than robust empirical evidence.

Already at the dawn of thalamic DBS for tremor in ET and PD, Benabid et al. reported in 1991, that a “beneficial effect on tremor began at more than 60 Hz, peaked from about 150 to 1000 Hz, and then slowly fell until 5000 Hz”, and in 1996 refined that “the optimum frequency was in the 100 Hz to 1000 Hz range with a minimum at approximately 250 Hz”.(7,8) In following studies, a sigmoidal dose–response relationship between tremor suppression and stimulation frequency with a linear improvement in tremor suppression between 0 Hz and 100 Hz and a flooring effect between 130 Hz and 185 Hz was established for thalamic DBS.(9–12)

In STN-DBS for PD, two landmark studies from the early 2000s, exploring a broad frequency range (5-250 Hz) in small cohorts (n = 10 and n = 12), demonstrated that frequencies above 50 Hz were necessary to significantly improve motor symptoms, whereas very low frequencies < 50 Hz exacerbated akinesia and rigidity.(13,14) No additional improvement in overall motor outcomes was observed beyond 130 Hz, leading to the establishment of 130 Hz as the clinical standard.(15) Moro et al. nonetheless reported a maximum effect on tremor suppression at 185 Hz compared with 130 Hz.(14)

However, these previous studies were limited to investigating a few patients(7,13) or examining the effects of combinations of stimulation parameters,(12,14,16) and did not account for differences in the total electrical energy delivered (TEED) to the tissue, which increases linearly with increasing stimulation frequency.(8,9,12,16) Importantly, the therapeutic effect and the occurrence of side-effects crucially depend on the applied TEED.(12,17) When calculating the applied TEED in the above-presented studies, there are indications that higher stimulation frequencies may achieve comparable or superior tremor control at lower energy delivery, implying a potentially greater stimulation efficiency.

Nevertheless, despite the absence of clear and robust data, a common programming routine has emerged over the past several years: in cases of insufficient tremor suppression, the stimulation frequency is typically increased from 130 Hz to 185 Hz. This empirical strategy is based largely on subjective clinical experience—often transmitted informally across generations of DBS programmers—rather than on systematic, data-driven evidence. As a result, current programming practices may reflect tradition and anecdotal expertise more than a validated data-driven rationale. This highlights the need for prospective studies aimed at defining evidence-based frequency optimization strategies for tremor control.

By isolating frequency effects under energy-equivalent conditions, the present study therefore aimed to clarify whether increasing DBS frequency beyond 130 Hz provides a therapeutic advantage for tremor suppression in VIM/PSA DBS for ET and STN-DBS for PD. We hypothesized that with TEED-equivalent stimulation, a higher stimulation frequency of 185 Hz would provide better tremor control.

## Methods

### Inclusion Criteria and Ethics

This study was a prospective single-center, double-blinded clinical trial. Patients aged ≥ 18 and ≤ 80 years with a confirmed diagnosis of ET or PD according to the International Parkinson and Movement Disorder Society consensus diagnostic criteria(18,19) and treatment with uni- or bilateral DBS in VIM/PSA (ET) or STN (PD) for at least three months were eligible for study participation. Only patients with a relevant tremor burden without stimulation (StimOFF) and after discontinuation of dopaminergic medication for at least 12 hours (MedOFF) in at least one upper extremity were included. For ET, a relevant tremor burden was defined as ≥ 2 of 4 points in one of the items 5 and 6 of the Fahn Tolosa Marin Tremor Rating Scale (FTM-TRS, postural or action/intention tremor)(20). For PD, a relevant tremor burden was defined as ≥ 2 of 4 points in item III.17 of the MDS-Unified Parkinson’s Disease rating scale (MDS-UPDRS III, rest tremor amplitude).(21) Only leads implanted in the hemisphere contralateral to a body side with relevant tremor burden were tested.

The local ethics committee of the University of Cologne approved the study (22–1435), which was conducted in accordance with the Declaration of Helsinki. It was registered with the German Clinical Trials Registry (DRKS00032470). All participants gave oral and written informed consent before participation. Clinical testing was conducted at the Department of Neurology of the University Hospital Cologne between 04/2023 and 08/2024.

### Study Design and Procedures

The study procedures are summarized in Figure 1. Clinical testing was conducted in the MedOFF condition. First, tremor control under previously established stimulation settings was evaluated. If necessary, contact choice was optimized clinically as per clinical routine at our center, before proceeding.

**Figure 1:**
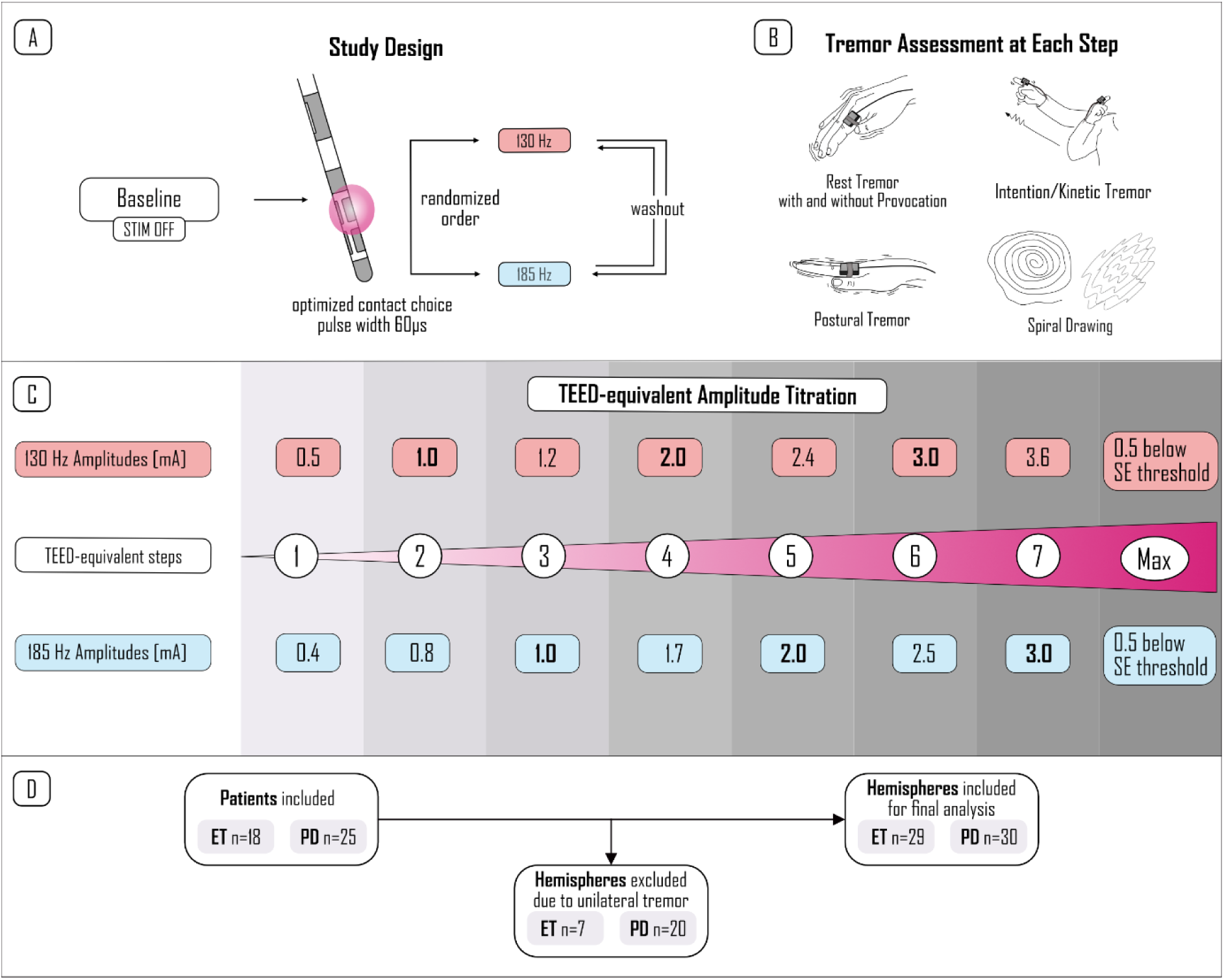
Study Design. (A) Study Design and procedures for one hemisphere, with a baseline assessment and two measurement series in randomized order regarding the frequency (130Hz or 185Hz). Pulse width (60µs) and contact choice were kept constant. (B) Tremor Assessment with each Stimulation Setting comprised Rest Tremor, Postural Tremor and Intention/Kinetic Tremor as well as spiral drawing. (C) Tremor Assessment was repeated at each TEED-equivalent Amplitude step. (D) Study flow chart. Abbreviations: ET = Essential Tremor, PD = Parkinson’s Disease, TEED = total electrical energy delivered, SE = side effect

Then, the overall stimulation effect on motor symptoms was evaluated by assessing the MDS-UPDRS III in PD and the FTM-TRS in ET in MedOFF/Stimulation ON (StimON) as well as after switching off stimulation for 30 minutes (“wash-out phase”) to reduce potential confounding from residual stimulation effects or rebound tremor.(22)

Before starting hemisphere-wise clinical testing, stimulation of the contralateral hemisphere was activated in the individual’s clinical settings to enhance patient comfort and to prevent measurement bias from contralateral tremor. First, a baseline assessment was performed. Then, two stimulation series were conducted, one at 130 Hz and one at 185 Hz, each comprising up to eight clinical assessments. The order of stimulation frequencies (130 Hz and 185 Hz) was randomized for each participant using balanced block randomization, and both the examiner and the patient were blinded to stimulation frequency (P.T.). Within each series, stimulation contacts, pulse width (60μs), and frequency were kept constant, while amplitude was gradually increased in TEED-equivalent steps. The chosen amplitudes ensured TEED comparability across frequencies and, simultaneously, enabled assessments at matched amplitudes (1, 2, and 3 mA; Figure 1C and Supplementary Table S1).

Clinical assessment included the evaluation of rest tremor (completely rested as well as with provocation by cognitively demanding such as calculating or spelling), postural, and kinetic tremor following the instructions of MDS-UPDRS III items III.15, III.16., and III.17, as well as spiral drawing with standardized video-based instructions (PsychToolbox Version 3.0.19 for MATLAB).(23,24) For kinetic tremor, participants were asked to press a button, reachable with the arm outstretched in front of them and to move the arm back to the wing position, alternatingly. Each task was performed for 20 seconds, with each trial’s beginning and end indicated by an acoustic signal. Tremor severity was measured by triaxial accelerometry at the index finger and wrist using a BrainProducts (BrainProducts GmbH, Gilching, Germany) triaxial accelerometer at the proximal dorsum of the index finger (Sampling rate 2500 Hz; ±2 g acceleration sensing range; <0.001 g resolution) and a triaxial wristwatch accelerometer (GENEActiv© Original, ActivinsightsTM, Kimbolton, UK; sampling rate 100 Hz; ±8 g acceleration sensing range; 0.004 g resolution). From the raw accelerometry data, an accelerometry tremor score was calculated using a previously validated approach (see Supplementary Material for Intention Tremor analysis).(2,25) For three hemispheres, the finger-worn accelerometer failed and data from the wristwatch accelerometer were used for further analysis. Additionally, tremor severity and spiral drawing were rated by an experienced clinician (CvdL), blinded to the current stimulation settings, using TETRAS item 6 and the TETRAS-provided tremor amplitude cutoffs. TETRAS was chosen as it provides smaller incremental steps (0.5-point scoring) than FTM-TRS, and has smaller ceiling effects.(26,27) In PD, akinetic-rigid symptoms were additionally evaluated based on MDS-UPDRS items III.3 (Upper Extremity Rigidity) and III.4 (Finger Tapping).

After each amplitude adjustment, a minimum of 60 seconds was waited before initiating the clinical assessments (“wash-in phase”). If persistent side effects occurred before the maximum amplitude was reached, the stimulation series was terminated, and the threshold for side effects was documented. If no side effects occurred after completing the series, amplitude was further increased until persistent side effects appeared (or up to 6 mA), and another tremor assessment was performed 0.5 mA below the frequency-specific side effect threshold for evaluating maximum possible efficacy.

Between the two stimulation series, a 10-30 minutes wash-out period was maintained until tremor severity returned to baseline to account for possible prolonged stimulation or rebound effects. For participants with bilateral tremor, the procedure was repeated for the contralateral hemisphere, with a minimum 30-minute pause between examinations of the second hemisphere.

Demographic information and individual imaging data for lead reconstructions were collected from the patients’ records. For visualization purposes, the DBS leads were reconstructed from postoperative CT scans and transformed into MNI (ICBM 2009b NLIN asym.) standard space utilizing a well-established workflow in the LEAD-DBS toolbox (v 3.2, www.lead-dbs.org).(28)

The individual TEED was calculated as the sum of the TEED of all active contacts calculated as TEED [µJ/s] = I² × f × pw × R (I = contact-wise current, f = frequency, pw = pulse width, R = contact-specific impedance). Missing impedance values (n = 1 hemisphere) were addressed with simple imputation (contact-wise) to preserve sample size.

### Outcomes

The primary endpoint was the difference in total tremor improvement relative to baseline with TEED-equivalent stimulation at 130 Hz and 185 Hz, measured by accelerometry. Total tremor was defined as the sum of subscores for rest tremor with provocation, postural, and kinetic tremor. Secondary outcomes included clinical tremor ratings, changes in tremor subscores, relative tremor improvement with maximal tolerated amplitude, and side-effect thresholds, as measured by amplitude and TEED. In ET, analyzed subscores included postural, intention tremor and spiral drawing. In PD, analyzed subscores included rest tremor, action tremor (sum of postural and intention tremor), spiral drawing and akinetic-rigid symptoms (sum of rigidity and finger tapping). Of note, subscores were evaluated only if the examined subscore was rated >0 at baseline.

#### Sample Size Calculation

Sample size was determined based on previous work investigating the effect of stimulation parameters on tremor control in PD and ET. As comparable studies on frequency-dependent tremor suppression were unavailable, the calculation was based on an estimated effect size of *d* = 0.64 from a prior study by our group.(29) Assuming a power of 0.9 and a two-sided alpha level of 0.05, the required sample size to detect differences in tremor improvement for 185 Hz versus 130 Hz was estimated as 28 hemispheres. Accounting for a potential dropout rate of 10%, a total of 30 hemispheres per target (VIM/PSA and STN) were planned, corresponding to approximately 15–20 people with essential tremor and 20–25 people with Parkinson’s disease.

### Statistical Analysis

All analyses were based on within-subject, repeated-measures data, collected at up to 7 TEED-equivalent stimulation steps for both frequencies (130 and 185 Hz). Primarily, we evaluated the frequency effect across TEED-steps on total tremor improvement, separately for PD and ET, using a linear mixed-effects model (LME) with fixed effects for stimulation frequency (185 Hz vs 130 Hz, individual within-subject centered TEED, their interaction (Frequency × TEED), and a random intercept for investigated hemisphere to account for within-subject correlation: *Tremor Improvement ∼ Frequency * TEED (within-subject centered) + (1 | investigated hemisphere).* Of note, sub-side-effect threshold measurements were not included in the LME.

Post-hoc, we conducted an exploratory pairwise comparison of tremor suppression at both frequencies at each TEED-equivalent as well as at each amplitude-equivalent step. After testing normality of paired differences (Shapiro-Wilk), Wilcoxon signed-rank tests were applied in all cases due to non-normality. Resulting p-values were adjusted using Benjamini-Hochberg FDR-correction. The same analysis was employed for secondary outcomes. The significance level was set to p < 0.05 for all analyses.

Given the negative superiority findings, we added an exploratory equivalence analysis to assess clinical similarity of frequencies at each TEED step and each amplitude step. The equivalence margin (± 20 percentage points of relative tremor improvement) was set in line with prior studies and applied to the 90% CIs of the bootstrapped median differences (10000 resamples).(2) Equivalence was concluded if the 90% CI lay within [−Δ, +Δ].

## Data Sharing

The main outcome data supporting the findings of this study are available via the open science framework (htt s://osf.io/) at xx (data added upon acceptance).

## Results

We included 18 people with ET (n = 11 with sufficient bilateral tremor), of which we excluded one from the primary analysis due to missing accelerometry data, resulting in 29 hemispheres with complete outcome data, and 25 people with PD (n = 5 with sufficient bilateral rest tremor), resulting in 30 hemispheres. Supplementary Table S2 shows demographic and clinical data of the study population. Contact settings and lead types are summarized in Supplementary Tables S3 and S4. Lead positions within the target regions (VIM/PSA for ET, STN in PD) were confirmed in all participants and are visualized in Supplementary Figure S1.

### Linear Mixed-Effect Models

In both ET and PD, the LME revealed a significant main effect of TEED, whereas neither the main effect of stimulation frequency nor the interaction between frequency and TEED reached significance (Table 1). Overall, these findings indicate that tremor suppression increased with higher energy delivery, but was not systematically modulated by stimulation frequency. For subscores and clinical rating, results were similar and are reported in detail in Supplementary Table S5.

**Table 1:** Results of the linear mixed-effect models for total tremor assessment. We report fixed-effect estimates (β), 95% CIs, two-sided p-values (alpha = 0.05) and the number of observations (N).Significant effects (p<0.05) are marked in bold. Abbreviations: TEED = total electrical energy delivered

| Fixed Effect | Estimate | 95% CI [lower; upper] | p-value | N |
| --- | --- | --- | --- | --- |
| **Essential Tremor** |  |  |  |  |
| **Intercept** | **0.625** | **0.555; 0.695** | **<0.001** | **300** |
| Frequency | -0.04 | -0.092; 0.012 | 0.135 | 300 |
| **TEED** | **0.007** | **0.005; 0.008** | **<0.001** | **300** |
| Frequency:TEED | 0 | -0.002; 0.002 | 0.892 | 300 |
| **Parkinson's Disease** |  |  |  |  |
| **Intercept** | **0.541** | **0.429; 0.653** | **<0.001** | **381** |
| Frequency | -0.026 | -0.083; 0.03 | 0.359 | 381 |
| **TEED** | **0.006** | **0.005; 0.008** | **<0.001** | **381** |
| Frequency:TEED | 0 | -0.002; 0.003 | 0.889 | 381 |

### Explorative Pairwise-Comparisons

#### Total Tremor

In ET, there were no differences in total tremor improvement between 130 Hz and 185 Hz stimulation, as assessed by accelerometry at neither when comparing each TEED-equivalent step nor when comparing amplitude-equivalent stimulation at 1 mA, 2 mA, and 3 mA (Figure 2A). Evaluated by clinical rating, stimulation at 185 Hz yielded significantly better tremor ratings at 1 mA (p = 0.003, FDR-corrected p = 0.009). However, this superiority was not persistent at higher amplitudes or under TEED-equivalent conditions (Supplementary Table S6A&B).

**Figure 2:**
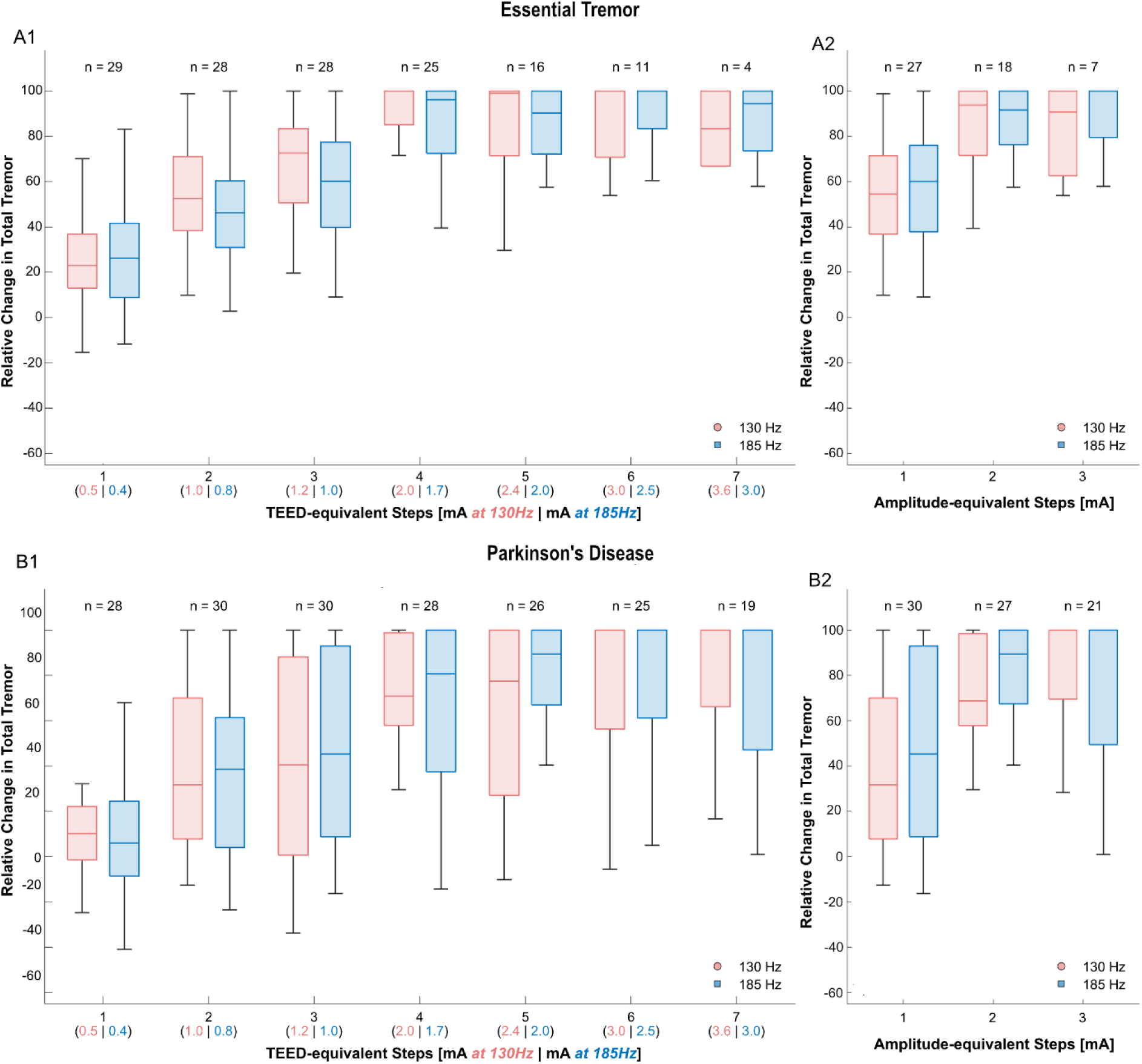
Effect of 130 Hz vs. **185 Hz DBS on Total Tremor Improvement** for ET (A) and PD (B), evaluated by accelerometry. Boxplots in panel A1 (ET) and B1 (PD) show total tremor improvement relative to baseline at each predefined TEED-equivalent step. Panel A2 (ET) and B2 (PD) show total tremor improvement with both stimulation frequency at 1, 2, and 3 mA. *p<0.05 after FDR-correction. Abbreviations: ET = Essential Tremor, PD = Parkinson’s Disease, TEED = total electrical energy delivered.

In PD, no significant frequency-related differences in total tremor control were detected under TEED-equivalent or amplitude-equivalent stimulation paradigms, neither by accelerometry nor clinical tremor ratings (Figure 2B, Supplementary Table S6A&B).

#### Tremor Subscores

In ET, there was a significant difference in postural tremor control as assessed by accelerometry favoring 130 Hz stimulation over 185 Hz during TEED-equivalent steps only at condition 3. However, this effect was not detectable in any other TEED-equivalent step or reproducible when assessed by clinical rating. For all other subscores no significant differences were detected under TEED-equivalent or amplitude-equivalent stimulation. (Supplementary Table S7).

For PD, there were no significant differences regarding tremor subscores. In contrast, for akinetic-rigid symptoms, stimulation at 185 Hz resulted in significantly better symptom control at 1 mA and 2 mA under amplitude-equivalent conditions; however, these differences were no longer significant after adjustment for TEED (Supplementary Table S8).

### Explorative Equivalence Analysis

Overall 90% CIs of the bootstrapped median differences of total tremor improvement measured by accelerometry in ET and PD lay within the equivalence margin (±20%), indicating overall equivalence. Of note, at TEED-equivalent step 7 in the ET cohort the 90% CI exceeded the equivalency margin in favor of 185Hz. However, there were only 4 observations in this group. Only at TEED-equivalent step 3 in ET the CI excluded zero and favored 130 Hz within the equivalency margin (Figure 3).

**Figure 3:**
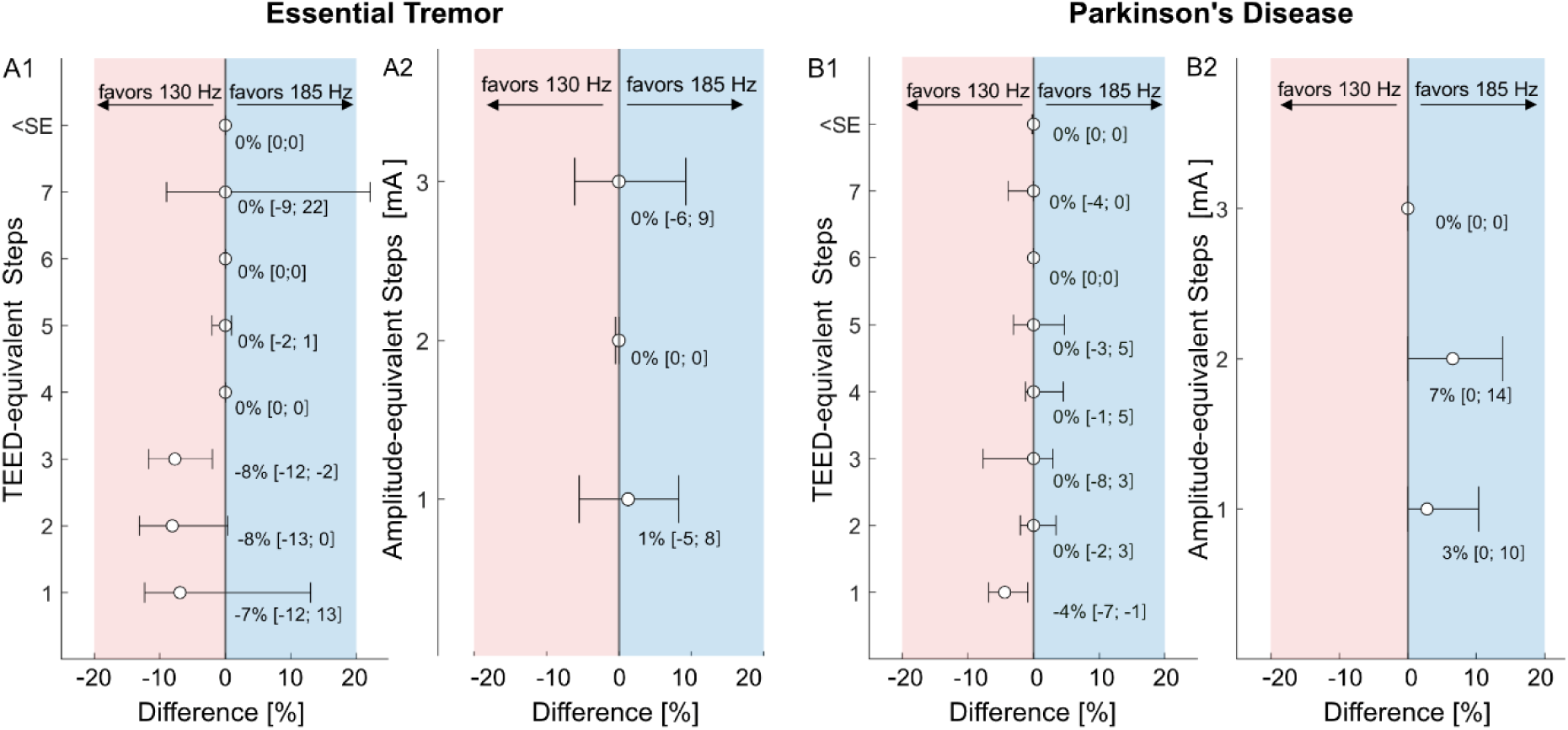
Explorative equivalence analysis of total tremor control with 130 Hz vs 185 Hz stimulation. Forest plots show bootstrapped median differences (185Hz – 130Hz frequency total tremor improvement) with 90% confidence intervals for TEED-equivalent (A1, B1) and amplitude-equivalent stimulation (A2, B2) in ET (A) and PD (B). Shaded areas represent the equivalence region (±20%). Negative values favor 130 Hz (red), positive values favor 185 Hz stimulation (blue).

### Side Effects

Results of comparisons at the side-effect threshold are illustrated in Figure 4 and Supplementary Table S8. In ET, maximum total tremor improvement reached ceiling levels, with a median tremor suppression of 100% and no significant difference between frequencies (p>1). Side-effect thresholds occurred at lower amplitudes for 185 Hz (2.5 mA [IQR 2.2-3.1]) than for 130 Hz (3.0 mA [IQR 2.5–3.5]; p = 0.001), but with comparable TEED values (79 µJ/s [39–116] vs 67 µJ/s [43–107]; p >1).

**Figure 4:**
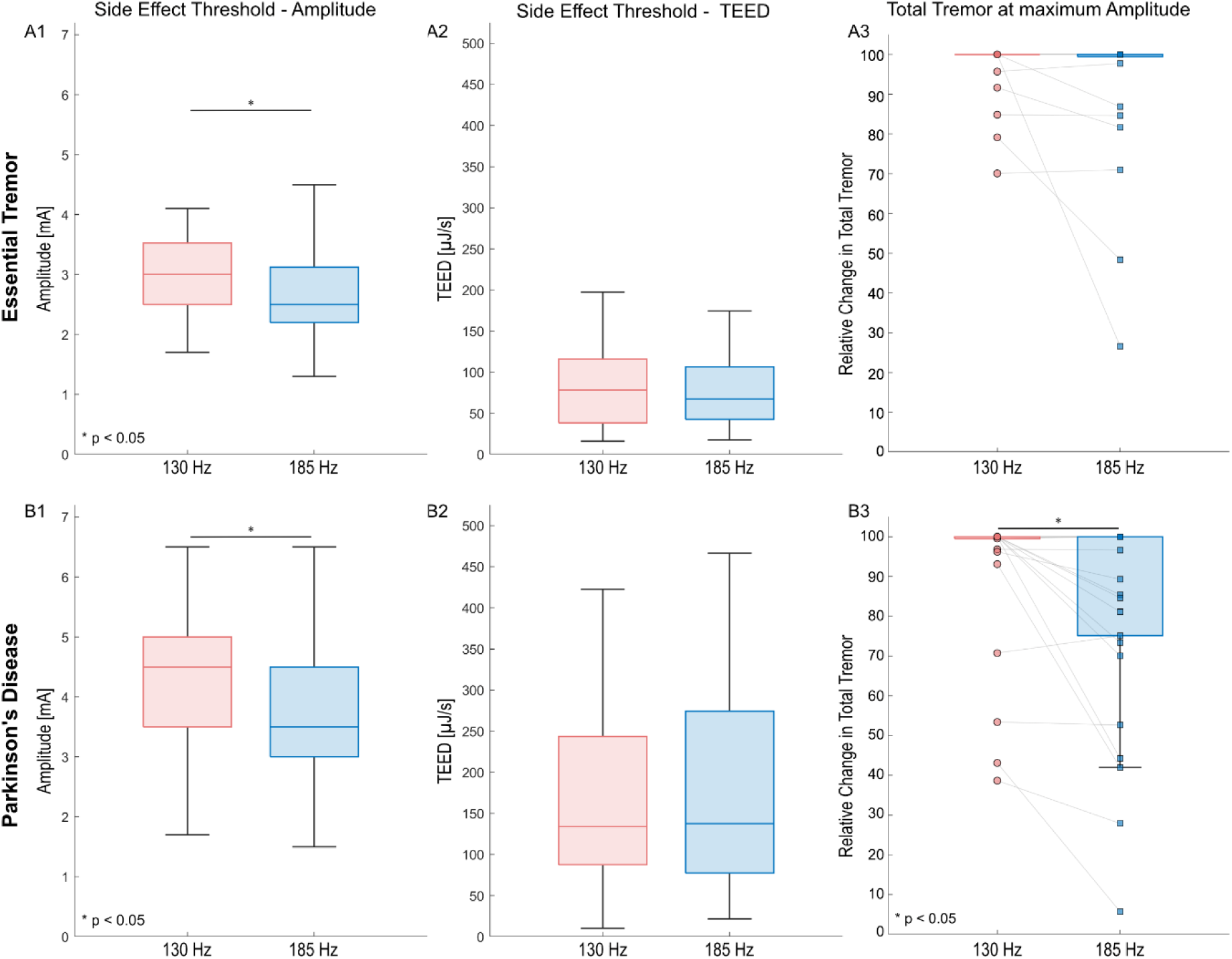
Side Effect Occurrence and Maximum Total Tremor Improvement. Boxplots show Amplitude [mA] (A1&B1) and TEED [µJ/s] (A2&B2) at the occurrence of persistent side-effects, as well as total tremor suppression 0.5 mA below side effect for stimulation with 130 Hz (red) and 185 Hz (blue), for ET (A) and PD (B).

In PD, although median tremor improvement <0.5 mA below the side effect threshold was 100% for both frequencies, variability was greater under 185 Hz compared to 130 Hz (130 Hz: 100% [IQR 96–100], 185 Hz: 100% [IQR 74–100]; p = 0.007, FDR-corrected p = 0.389). Side-effect thresholds were again lower for 185 Hz (3.5 mA [3.0–4.5] than for 130 Hz (4.5 mA [IQR 3.5–5.0]; p < 0.001), while TEED values at the side-effect threshold did not differ significantly (134 µJ/s [88–243] vs 137 µJ/s [78–274]; p > 1).

## Discussion

This is the first prospective study systematically comparing 130 Hz and 185 Hz stimulation for tremor suppression in VIM/PSA-DBS for ET and STN-DBS for PD, applying TEED-equivalent stimulation to control for differences in delivered stimulation energy when varying stimulation frequencies. Across all target points and tremor subscores, no consistent superiority of either frequency could be demonstrated when applying TEED-equivalent stimulation. Linear mixed-effects models and non-parametric pairwise comparisons revealed comparable tremor control at 130 Hz and 185 Hz, and the exploratory equivalence analysis confirmed clinical equivalency within a ±20 % margin of relative tremor improvement. This study thereby provides evidence that there is currently no rationale for routinely investigating DBS above 130 Hz for tremor suppression.

As outlined in the introduction, the absence of any consistent clinical benefit from 185 Hz stimulation aligns with earlier studies reporting that tremor improvement plateaus at approximately 100–130 Hz in both PD and ET. (9,11–14,16) From a biophysical perspective, this frequency range likely marks the threshold at which axonal firing within the stimulated volume becomes fully “overwritten”, thereby disrupting pathological oscillations within the cerebello-thalamo-cortical or basal ganglia-thalamo-cortical loops. Further increases in frequency beyond this saturation point might not recruit additional axons or enhance desynchronization but merely raise the number of delivered pulses and, consequently, the total energy consumption.(30,31)

The present study underscores a strong relationship between TEED and tremor improvement as shown by a saturation curve with a clear ceiling effect (Figure 2). In contrast to previous studies investigating the effect of varying stimulation frequencies we meticulously accounted for TEED in our analysis. As TEED is a non-linear function of amplitude, pulse width, and frequency, this is of utmost importance when comparing the effect of varying stimulation parameters. Recent studies have demonstrated that neglecting TEED yields to controversial results, e.g., when investigating short pulse widths or low-frequency stimulation.(32–35) This is also represented in our data with 185Hz stimulation resulting in superior improvement in akinetic-rigid symptoms when comparing stimulations at equal amplitudes of 1 mA or 2 mA in PD, but not when comparing TEED-equivalent stimulation (Supplementary Table S7).

Given the strong relationship between tremor improvement and TEED, the clinician may consider increasing the stimulation frequency from 130Hz to 185Hz could be considered to improve tremor control when increasing the amplitude is limited by side-effects. Assuming a stimulation with 1mA and a stand impedance of 1 kΩ this approach increases the TEED by 42.3%, resembling an increase of the amplitude by 0.2 mA. However, when comparing amplitude-equivalent stimulation settings at 1 mA, 2 mA and 3 mA there was no difference in tremor control on the group level. In line with Kuncel et al.(16), amplitude-wise side-effect thresholds were even lower during stimulation was performed at 185 Hz throughout the present study. Additionally, increasing stimulation frequency might increase local stimulation intensity and thereby increase the likelihood of stimulation-induced side effects such as ataxia or lead to habitua84tion by supramaximal stimulation.(36,37) This is supported by the fact that tremor improvement at 0.5 mA below the side-effect threshold was significantly better at 130 Hz than at 185 Hz in PD, suggesting that the lower frequency may provide a more favorable balance of effects and side effects. Of note, there were no differences in side-effect thresholds when TEED was accounted for (Figure 4).

### Limitations

Several limitations must be considered. First, the study was originally powered for superiority testing, not specifically for equivalence; although the equivalence analysis was exploratory, its consistent results across models support the clinical equivalence of the two frequencies. Second, TEED is a composite measure that includes stimulation frequency as one of its factors. Consequently, modeling both TEED and frequency within the same model can introduce partial collinearity, potentially complicating the interpretation of independent parameter effects. However, despite this shared variance, frequency did not show a significant main effect, nor did it interact with TEED. If frequency exerted a clinically meaningful effect beyond its contribution to total electrical energy, such an effect would be expected to emerge even in the presence of collinearity. Third, analyses were restricted to acute stimulation sessions. Therefore, long-term effects, including habituation, or adaptation phenomena cannot be inferred.

## Conclusion

This study provides Level II evidence that a higher stimulation frequency of 185 Hz does not offer additional benefit in deep brain stimulation for tremor and supports 130 Hz as the standard stimulation frequency for tremor suppression in ET and PD.

## Data Availability

The main outcome data supporting the findings of this study are available via the open science framework (data added upon acceptance in peer-reviewed Journal). All data produced in the present study are available upon reasonable request to the authors.

## Acknowledgements

We thank patients and their caregivers for their participation and commitment. We are grateful to the healthcare professionals who make complex care possible - especially Study Nurses Max Pohl and Justus Rewolle, and Parkinson Nurse Susanne Hoffmann.

## Authors’ Roles

(1) Research project: A. Conception, B. Organization, C. Execution; (2) Statistical analysis: A. Design, B. Execution, C. Review and critique; (3) Manuscript: A. Writing of the first draft, B. Review and critique.

C.v.d.L.: 1A, 1B, 1C, 2A, 2B, 3A

P.T.: 1B, 1C, 2B, 2C, 3B

T.A.D.: 1A, 2A, 2C, 3B

C.S.-T.:1C, 2C,3B

G.A.B.: 2A, 2C, 3B

H.J.: 1A, 2C, 3B

G.R.F.: 1B, 2C, 3B

V.V.-V.:2C, 3B

M.T.B.: 1A, 1B, 2C, 3B

J.N.P.-S.:1A, 1B, 2A, 2C, 3A

## Supplementary Material

### Accelerometry Model Parameters for Intention/Kinetic Tremor

Accelerometric assessment of intention tremor mainly followed the previously published Method in van der Linden et al. 2023 and 2026.^1,2^ The previously published model resulted in a numerical good fit for Intention Tremor (Finger [Wrist]: weighted kappa = 0.5 [0.4], percentage concordance of 67.4% [63.1%]). However, it resulted in a nearly linear relationship between mean Acceleration and predicted tremor score due to the low number of observations with higher clinical ratings. Therefor the model was adapted by weighting each value of clinical rating (0,1,2,3,4) by the number of observations. Numerically the new model resulted in lower weighted kappa and percentage concordance at lower clinical ratings, but a higher concordance at higher clinical ratings.

**Supplementary Figure:**
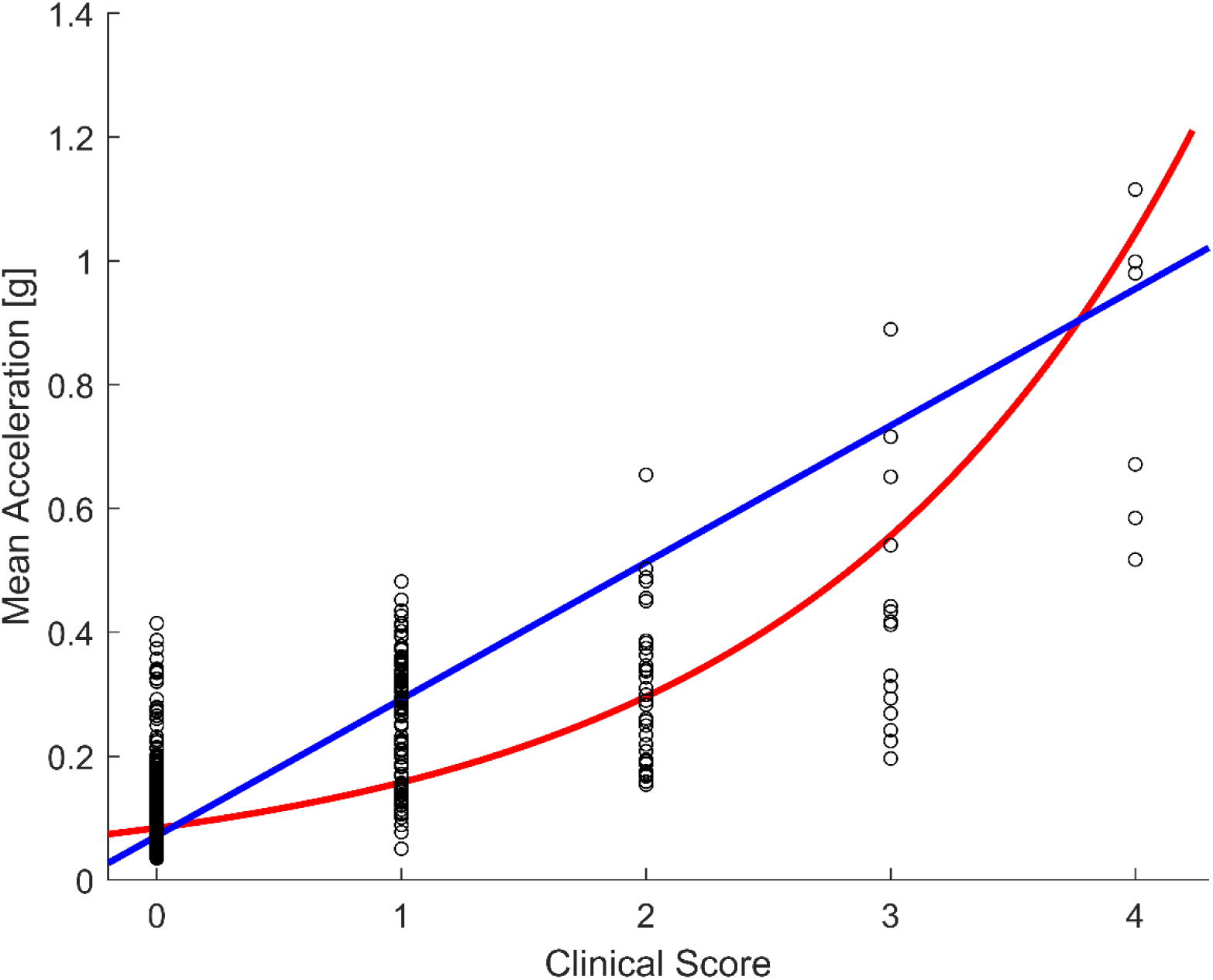
Illustration of the fit of the non-weighted (blue) and the weighted model (red) for accelerometric intention tremor ratings by finger accelerometry.

**Supplementary Table:** Accelerometry Model Parameters for Intention/Kinetic Tremor. Model parameters (coefficients a, b, and c) determined in the training dataset, weighted Kappa and Concordance between clinical ratings and the resulting accelerometric tremor score when applying the model parameters to the validation data set. Abbreviations: *a* = slope, *b* = intercept, *c* = exponent of a power relationship between *T* and *TR*, RMSE = root mean squared error, T = tremor severity assessed by Mean Acceleration, TR = tremor rating score

| Model | Location | TR = a * T<sup>c</sup> + b |  |  | RMSE | Weighted Kappa | Concordance |
| --- | --- | --- | --- | --- | --- | --- | --- |
|  |  | a | b | c |  |  |  |
| non-weighted | Finger | 4.47 | -0.39 | 0.94 | 0.628 | 0.50 | 0.674 |
|  | Wrist | 8.91 | -0.29 | 1.11 | 0.917 | 0.40 | 0.631 |
| weighted | Finger | 10.6 | -6.6 | 0.16 | 0.085 | 0.15 | 0.409 |
|  | Wrist | 209.17 | -201.76 | 0.05 | 0.111 | 0.16 | 0.249 |

**Supplementary Table S1:** Amplitudes of TEED-equivalent Steps. Amplitudes of the two measurement series for ET (A for 130 Hz, B for 185 Hz; differences in TEED values result from rounding).

| | Amplitudes 130 Hz<br>[mA] | Amplitudes 185 Hz<br>[mA] | TEED [μJ/s] (1000<br>Ohm, 60 μs, 130<br>Hz) | TEED [μJ/s] (1000<br>Ohm, 60 μs, 185Hz<br>) |
| --- | --- | --- | --- | --- |
| **1** | 0.5 | 0.4 | 2 | 1.8 |
| **2** | 1 | 0.8 | 7.8 | 7.1 |
| **3** | 1.2 | 1 | 11.2 | 11.1 |
| **4** | 2 | 1.7 | 31.2 | 32.1 |
| **5** | 2.4 | 2 | 44.9 | 44.4 |
| **6** | 3 | 2.5 | 70.2 | 69.4 |
| **7** | 3.6 | 3 | 101.1 | 99.9 |

**Supplementary Table S2:**
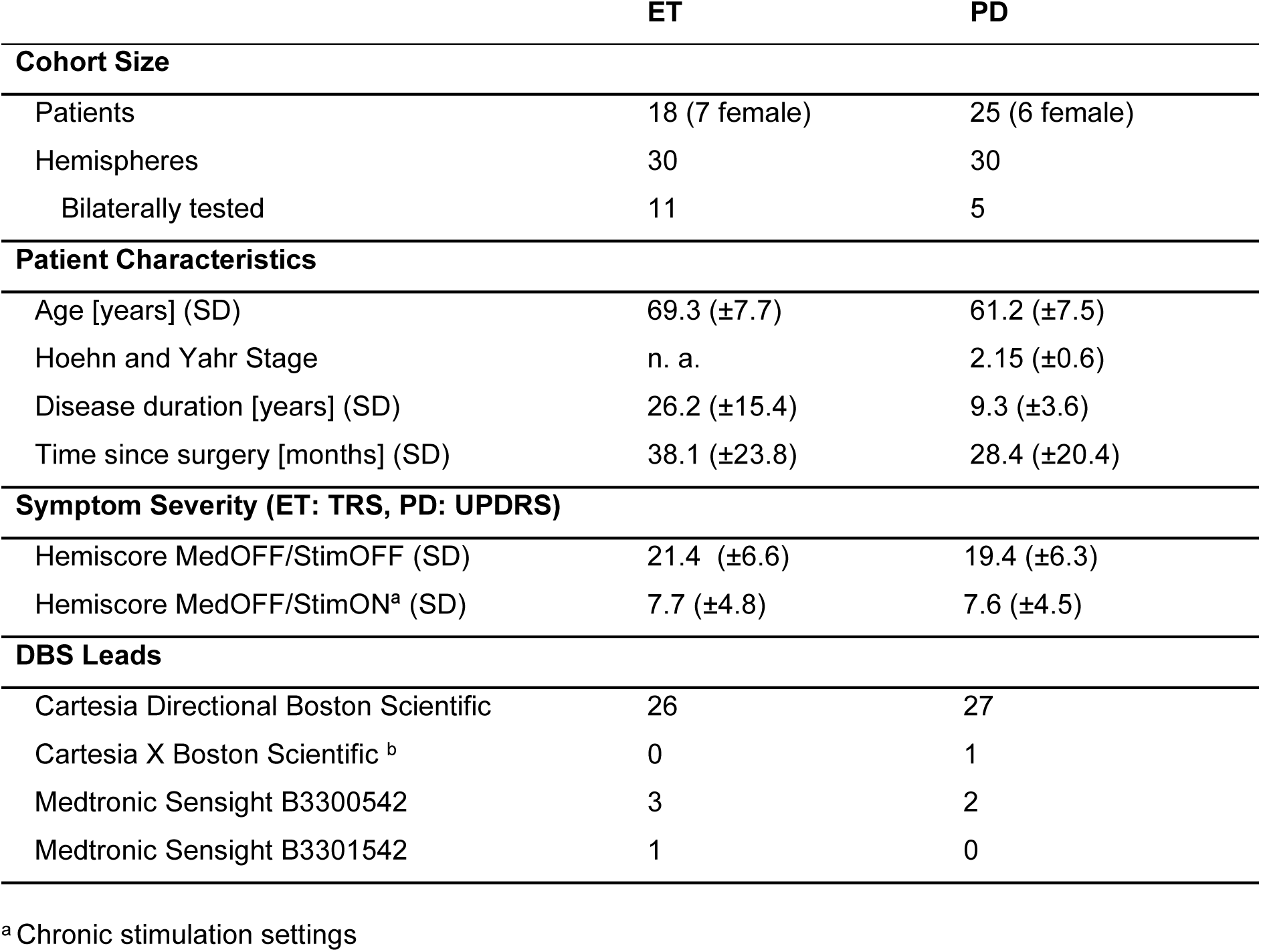
Patient Characteristics. Abbreviations: DBS = deep brain stimulation, ET = Essential Tremor, MedOFF = after 12 hours withdrawal of disease-specific medication, PD = Parkinson’s Disease, SD = standard deviation, StimOFF = Stimulation OFF, StimON = Stimulation ON, TRS = Fahn Tolosa Marin Tremor Rating Scale, UPDRS = Unified Parkinson’s Disease Rating Scale

**Supplementary Table S3:** Contact Settings of the Essential Tremor Cohort. Contact labeling was simplified to enable comparability across different lead types with contact 1 representing the most ventral and 8 the most dorsal contact. 2/3/4 and 5/6/7 correspond to the directional segments A/B/C counter-clockwise.

| 1[%] | 2[%] | 3[%] | 4[%] | 5[%] | 6[%] | 7[%] | 8[%] | Hemisphere | Lead Type |
| --- | --- | --- | --- | --- | --- | --- | --- | --- | --- |
| 0 | -34 | -33 | -33 | 0 | 0 | 0 | 0 | right | Vercise Cartesia |
| 0 | -100 | 0 | 0 | 0 | 0 | 0 | 0 | right | Vercise Cartesia |
| -100 | 0 | 0 | 0 | 0 | 0 | 0 | 0 | left | Vercise Cartesia |
| 0 | 0 | 0 | 0 | 0 | -80 | -60 | 0 | right | Vercise Cartesia |
| 0 | -50 | -50 | 0 | 0 | 0 | 0 | 0 | left | Vercise Cartesia |
| 0 | -34 | -33 | -33 | 0 | 0 | 0 | 0 | right | Vercise Cartesia |
| 0 | -34 | -33 | -33 | 0 | 0 | 0 | 0 | left | Sensight B3300542 |
| -50 | 0 | 0 | -50 | 0 | 0 | 0 | 0 | right | Vercise Cartesia |
| 0 | 0 | 0 | -80 | 0 | 0 | -20 | 0 | left | Vercise Cartesia |
| 0 | 0 | 0 | 0 | -70 | 0 | -30 | 0 | right | Vercise Cartesia |
| 0 | 0 | -25 | -75 | 0 | 0 | 0 | 0 | left | Vercise Cartesia |
| 0 | 0 | -50 | -50 | 0 | 0 | 0 | 0 | right | Vercise Cartesia |
| -20 | -20 | 0 | -60 | 0 | 0 | 0 | 0 | left | Vercise Cartesia |
| -40 | 0 | 0 | -60 | 0 | 0 | 0 | 0 | right | Vercise Cartesia |
| -100 | 0 | 0 | 0 | 0 | 0 | 0 | 0 | left | Vercise Cartesia |
| -100 | 0 | 0 | 0 | 0 | 0 | 0 | 0 | right | Vercise Cartesia |
| 0 | 0 | -100 | 0 | 0 | 0 | 0 | 0 | right | Vercise Cartesia |
| 0 | 0 | -100 | 0 | 0 | 0 | 0 | 0 | left | Vercise Cartesia |
| -100 | 0 | 0 | 0 | 0 | 0 | 0 | 0 | left | Vercise Cartesia |
| -60 | -14 | -13 | -13 | 0 | 0 | 0 | 0 | right | Vercise Cartesia |
| 0 | -34 | -33 | -33 | 0 | 0 | 0 | 0 | left | Sensight B3301542 |
| 0 | 0 | 0 | 0 | 0 | -40 | -60 | 0 | left | Sensight B3300542 |
| 0 | -34 | -33 | -33 | 0 | 0 | 0 | 0 | right | Sensight B3300542 |
| -100 | 0 | 0 | 0 | 0 | 0 | 0 | 0 | left | Vercise Cartesia |
| -100 | 0 | 0 | 0 | 0 | 0 | 0 | 0 | right | Vercise Cartesia |
| 0 | -34 | -33 | -33 | 0 | 0 | 0 | 0 | right | Vercise Cartesia |
| 0 | -34 | -33 | -33 | 0 | 0 | 0 | 0 | left | Vercise Cartesia |
| 0 | 0 | 0 | 0 | -10 | -80 | -10 | 0 | right | Vercise Cartesia |
| 0 | -34 | -33 | -33 | 0 | 0 | 0 | 0 | left | Vercise Cartesia |
| 0 | -24 | -23 | -23 | -10 | -10 | -10 | 0 | right | Vercise Cartesia |

**Supplementary Table S4:** Contact Settings of the Parkinson’s Disease Cohort. Contact labeling was simplified to enable comparability across different lead types with contact 1 representing the most ventral and 8 the most dorsal contact. 2/3/4 and 5/6/7 correspond to the directional segments A/B/C counter-clockwise.

| 1[%] | 2[%] | 3[%] | 4[%] | 5[%] | 6[%] | 7[%] | 8[%] | Hemisphere | Lead Type |
| --- | --- | --- | --- | --- | --- | --- | --- | --- | --- |
| 0 | 0 | 0 | -50 | 0 | 0 | -50 | 0 | right | Vercise Cartesia |
| 0 | 0 | 0 | 0 | 0 | -50 | 0 | -50 | left | Vercise Cartesia |
| 0 | 0 | 0 | 0 | -34 | -33 | -33 | 0 | left | Vercise Cartesia |
| 0 | 0 | -50 | 0 | 0 | -50 | 0 | 0 | left | Vercise Cartesia |
| 0 | 0 | 0 | 0 | -34 | -33 | -33 | 0 | left | Vercise Cartesia |
| 0 | 0 | 0 | 0 | 0 | -35 | -35 | -30 | right | Vercise Cartesia |
| 0 | 0 | 0 | 0 | 0 | -75 | -25 | 0 | right | Vercise Cartesia X |
| 0 | -10 | -10 | -10 | -23 | -23 | -24 | 0 | left | Vercise Cartesia |
| 0 | 0 | -70 | 0 | 0 | -30 | 0 | 0 | right | Vercise Cartesia |
| 0 | 0 | 0 | 0 | -34 | -33 | -33 | 0 | left | Vercise Cartesia |
| 0 | -10 | -10 | -10 | -22 | -24 | -24 | 0 | right | Vercise Cartesia |
| 0 | 0 | 0 | 0 | 0 | 0 | -40 | -60 | left | Vercise Cartesia |
| 0 | 0 | -30 | -30 | 0 | -20 | -20 | 0 | right | Vercise Cartesia |
| 0 | 0 | 0 | 0 | -100 | 0 | 0 | 0 | left | Vercise Cartesia |
| 0 | 0 | 0 | 0 | -50 | -50 | 0 | 0 | right | Vercise Cartesia |
| -40 | 0 | -15 | -45 | 0 | 0 | 0 | 0 | left | Vercise Cartesia |
| 0 | 0 | 0 | 0 | -24 | -23 | -23 | -30 | left | Vercise Cartesia |
| 0 | 0 | 0 | 0 | -34 | -33 | -33 | 0 | right | Vercise Cartesia |
| 0 | 0 | 0 | 0 | -34 | -33 | -33 | 0 | left | Vercise Cartesia |
| 0 | 0 | 0 | 0 | -30 | 0 | -30 | -40 | right | Vercise Cartesia |
| 0 | 0 | 0 | 0 | -100 | 0 | 0 | 0 | left | Vercise Cartesia |
| 0 | 0 | 0 | 0 | 0 | -25 | -75 | 0 | left | Vercise Cartesia |
| 0 | 0 | 0 | 0 | 0 | 0 | -40 | -60 | left | Vercise Cartesia |
| 0 | 0 | 0 | 0 | 0 | -25 | -75 | 0 | left | Vercise Cartesia |
| 0 | 0 | 0 | 0 | 0 | -25 | -75 | 0 | right | Vercise Cartesia |
| 0 | -50 | 0 | 0 | -50 | 0 | 0 | 0 | right | Sensight B3300542 |
| 0 | 0 | -100 | 0 | 0 | 0 | 0 | 0 | right | Vercise Cartesia |
| 0 | 0 | 0 | 0 | -75 | 0 | -25 | 0 | left | Vercise Cartesia |
| 0 | 0 | 0 | 0 | -100 | 0 | 0 | 0 | right | Vercise Cartesia |
| 0 | -34 | -33 | -33 | 0 | 0 | 0 | 0 | left | Sensight B3300542 |

**Supplementary Table S5:** Linear Mixed Model Results (Secondary Outcomes). Results of the linear mixed effect models for tremor subscores, rated clinically and evaluated by accelerometery. We report fixed-effect estimates (β), 95% CIs, two-sided p-values (alpha = 0.05) and the number of observations (N). Abbreviations: ACC = accelerometry.

| Outcome | Fixed Effect | Estimate | 95% CI<br>[lower; upper] | p-value<br>(uncorr.) | p-value<br>(FDR) | N |
| --- | --- | --- | --- | --- | --- | --- |
| **Essential Tremor** |  |  |  |  |  |  |
| Postural Tremor [ACC] | **Intercept** | **0.66** | **0.581; 0.74** | **<0.001** | **<0.001** | **300** |
|  | Frequency | -0.06 | -0.126; 0.006 | 0.073 | 0.499 | 300 |
|  | **TEED** | **0.005** | **0.004; 0.007** | **<0.001** | **<0.001** | **300** |
|  | Frequency:TEED | 0.001 | -0.001; 0.004 | 0.388 | >1 | 300 |
| Intention Tremor [ACC] | **Intercept** | **0.592** | **0.505; 0.678** | **<0.001** | **<0.001** | **300** |
|  | Frequency | -0.028 | -0.089; 0.033 | 0.374 | >1 | 300 |
|  | **TEED** | **0.007** | **0.006; 0.009** | **<0.001** | **<0.001** | **300** |
|  | Frequency:TEED | 0 | -0.003; 0.002 | 0.841 | >1 | 300 |
| Postural Tremor [Clinical] | **Intercept** | **0.67** | **0.605; 0.736** | **<0.001** | **<0.001** | **301** |
|  | Frequency | -0.053 | -0.114; 0.009 | 0.095 | 0.499 | 301 |
|  | **TEED** | **0.006** | **0.005; 0.008** | **<0.001** | **<0.001** | **301** |
|  | Frequency:TEED | <0.001 | -0.002; 0.003 | 0.752 | >1 | 301 |
| Intention Tremor [Clinical] | **Intercept** | **0.658** | **0.591; 0.725** | **<0.001** | **<0.001** | **301** |
|  | Frequency | -0.04 | -0.093; 0.012 | 0.131 | 0.499 | 301 |
|  | **TEED** | **0.007** | **0.006; 0.008** | **<0.001** | **<0.001** | **301** |
|  | Frequency:TEED | <0.001 | -0.002; 0.002 | 0.857 | >1 | 301 |
| Spiral [Clinical] | **Intercept** | **0.76** | **0.511; 0.64** | **<0.001** | **<0.001** | **301** |
|  | Frequency | <0.001 | -0.047; 0.047 | 0.994 | >1 | 301 |
|  | **TEED** | **0.006** | **0.005; 0.008** | **<0.001** | **<0.001** | **301** |
|  | Frequency:TEED | <0.001 | -0.002; 0.002 | 0.927 | >1 | 301 |
| **Parkinson's Disease** |  |  |  |  |  |  |
| Resting Tremor [ACC] | **Intercept** | **0.588** | **0.483; 0.692** | **<0.001** | **<0.001** | **381** |
|  | Frequency | -0.008 | -0.073; 0.057 | 0.808 | >1 | 381 |
|  | **TEED** | **0.006** | **0.005; 0.008** | **<0.001** | **<0.001** | **381** |
|  | Frequency:TEED | 0 | -0.002; 0.001 | 0.762 | >1 | 381 |
| Resting Tremor Provocation [ACC] | **Intercept** | **0.528** | **0.425; 0.63** | **<0.001** | **<0.001** | **381** |
|  | Frequency | 0.001 | -0.06; 0.062 | 0.972 | >1 | 381 |
|  | **TEED** | **0.006** | **0.005; 0.007** | **<0.001** | **<0.001** | **381** |
|  | Frequency:TEED | <0.001 | -0.002; 0.001 | 0.752 | >1 | 381 |
| Action Tremor [ACC] | **Intercept** | **0.502** | **0.353; 0.651** | **<0.001** | **<0.001** | **278** |
|  | Frequency | -0.036 | -0.105; 0.033 | 0.305 | >1 | 278 |
|  | **TEED** | **0.005** | **0.004; 0.006** | **<0.001** | **<0.001** | **278** |
|  | Frequency:TEED | 0.001 | -0.001; 0.002 | 0.55 | >1 | 278 |
| Resting Tremor [Clinical] | **Intercept** | **0.603** | **0.502; 0.704** | **<0.001** | **<0.001** | **382** |
|  | Frequency | -0.024 | -0.096; 0.048 | 0.518 | >1 | 382 |
|  | **TEED** | **0.006** | **0.005; 0.008** | **<0.001** | **<0.001** | **382** |
|  | Frequency:TEED | <0.001 | -0.002; 0.002 | 0.906 | >1 | 382 |
| Resting Tremor Provocation [Clinical] | **Intercept** | **0.546** | **0.454; 0.639** | **<0.001** | **<0.001** | **382** |
|  | Frequency | 0.008 | -0.051; 0.067 | 0.786 | >1 | 382 |
|  | **TEED** | **0.006** | **0.005; 0.007** | **<0.001** | **<0.001** | **382** |
|  | Frequency:TEED | <0.001 | -0.002; 0.001 | 0.855 | >1 | 382 |
| Action Tremor [Clinical] | **Intercept** | **0.578** | **0.466; 0.691** | **<0.001** | **<0.001** | **278** |
|  | Frequency | -0.023 | -0.087; 0.041 | 0.476 | >1 | 278 |
|  | **TEED** | **0.005** | **0.004; 0.006** | **<0.001** | **<0.001** | **278** |
|  | Frequency:TEED | <0.001 | -0.001; 0.002 | 0.791 | >1 | 278 |
| Spiral [Clinical] | **Intercept** | **0.715** | **0.586; 0.844** | **<0.001** | **<0.001** | **243** |
|  | Frequency | -0.02 | -0.103; 0.063 | 0.632 | >1 | 243 |
|  | **TEED** | **0.003** | **0.001; 0.004** | **<0.001** | **<0.001** | **243** |
|  | Frequency:TEED | <0.001 | -0.002; 0.002 | 0.746 | >1 | 243 |
| Akinetic-rigid Symptoms | **Intercept** | **0.562** | **0.497; 0.627** | **<0.001** | **<0.001** | **382** |
|  | Frequency | 0.021 | -0.024; 0.067 | 0.359 | >1 | 382 |
|  | **TEED** | **0.006** | **0.005; 0.007** | **<0.001** | **<0.001** | **382** |
|  | Frequency:TEED | -0.001 | -0.002; 0.001 | 0.299 | >1 | 382 |

**Supplementary Table S6A:** Pairwise Comparisons – Total Tremor.

|  | Comparison | Median Improvement<br>130 Hz [IQR] in % | Median<br>Improvement<br>185 Hz [IQR] in % | p-Value<br>(uncorr.) | p-Value<br>(FDR) | N |
| --- | --- | --- | --- | --- | --- | --- |
| **ET** | **TEED-Steps** |  |  |  |  |  |
|  | 1 | 23.1 [13-36.8] | 26.2 [8.9-41.6] | 0.854 | >1 | 29 |
|  | 2 | 52.6 [38.4-71.1] | 46.3 [31.1-60.4] | 0.151 | >1 | 28 |
|  | 3 | 72.7 [50.6-83.4] | 60.2 [39.8-77.5] | 0.003 | 0.064 | 28 |
|  | 4 | 100 [85.1-100] | 96.1 [72.5-100] | 0.583 | >1 | 25 |
|  | 5 | 99.1 [71.5-100] | 90.3 [72.1-100] | 0.365 | >1 | 16 |
|  | 6 | 100 [70.8-100] | 100 [83.4-100] | 0.813 | >1 | 11 |
|  | 7 | 23.1 [13-36.8] | 26.2 [8.9-41.6] | 0.854 | >1 | 29 |
|  | **Amplitudes** |  |  |  |  |  |
|  | 1 mA | 54.6 [36.8-71.5] | 60.1 [37.9-76] | 0.755 | >1 | 27 |
|  | 2 mA | 93.8 [71.6-100] | 91.7 [76.2-100] | 0.898 | >1 | 18 |
|  | 3 mA | 90.8 [62.6-100] | 100 [79.5-100] | 0.25 | >1 | 7 |
| **PD** | **TEED-Steps** |  |  |  |  |  |
|  | 1 | 10.1 [-1.5-22.2] | 6 [-8.6-24.4] | 0.055 | 0.594 | 28 |
|  | 2 | 31.6 [7.8-70.1] | 38.6 [4-61.3] | 0.909 | >1 | 30 |
|  | 3 | 40.5 [0.7-88.1] | 45.3 [8.8-93] | 0.925 | >1 | 30 |
|  | 4 | 70.8 [57.9-98.8] | 80.8 [37.5-100] | 0.831 | >1 | 28 |
|  | 5 | 77.4 [27.1-100] | 89.5 [66.9-100] | 0.911 | >1 | 26 |
|  | 6 | 100 [56.4-100] | 100 [61.2-100] | 0.455 | >1 | 25 |
|  | 7 | 100 [66.2-100] | 100 [47.1-100] | 0.195 | >1 | 19 |
|  | **Amplitudes** |  |  |  |  |  |
|  | 1 mA | 31.6 [7.8-70.1] | 45.3 [8.8-93] | 0.292 | 0.803 | 30 |
|  | 2 mA | 68.7 [57.8-98.4] | 89.5 [67.5-100] | 0.045 | 0.246 | 27 |
|  | 3 mA | 100 [69.6-100] | 100 [49.5-100] | 0.557 | >1 | 21 |

**Supplementary Table S6B:** Pairwise Comparisons – Total Tremor Clinical Rating.

|  | Comparison | Median Improvement<br>130 Hz [IQR] in % | Median Improvement<br>185 Hz [IQR] in % | p-Value<br>(uncorr.) | p-Value<br>(FDR) | N |
| --- | --- | --- | --- | --- | --- | --- |
| ET | **TEED-Steps** |  |  |  |  |  |
|  | 1 | 30 [14.3-39.6] | 26.7 [14.8-37] | 0.563 | >1 | 29 |
|  | 2 | 57.1 [41.7-68.8] | 55 [36.5-64.9] | 0.19 | >1 | 29 |
|  | 3 | 73.6 [62.7-80.6] | 64.6 [52.1-80] | 0.033 | 0.726 | 28 |
|  | 4 | 92.9 [71.9-97.3] | 93.3 [76-100] | 0.988 | >1 | 25 |
|  | 5 | 92.3 [73.9-93.8] | 87.5 [77.1-94.4] | 0.637 | >1 | 16 |
|  | 6 | 92.9 [82.5-95.8] | 95 [85.6-96.5] | 0.43 | >1 | 11 |
|  | 7 | 86.4 [71.8-96.1] | 84.2 [71-95.8] | 0.875 | >1 | 4 |
|  | **Amplitudes** |  |  |  |  |  |
|  | 1 mA | **57.1 [41.7-66.7]** | **64.6 [52.1-80]** | **0.003** | **0.016** | **28** |
|  | 2 mA | 91.4 [62.5-95] | 88.8 [79.2-96.4] | 0.055 | 0.15 | 18 |
|  | 3 mA | 91.7 [71.9-95.6] | 91.7 [78.8-98.8] | 0.188 | 0.344 | 7 |
| PD | **TEED-Steps** |  |  |  |  |  |
|  | 1 | 10.1 [-1.5-22.2] | 6 [-8.6-24.4] | 0.055 | 0.594 | 28 |
|  | 2 | 31.6 [7.8-70.1] | 38.6 [4-61.3] | 0.909 | >1 | 30 |
|  | 3 | 40.5 [0.7-88.1] | 45.3 [8.8-93] | 0.925 | >1 | 30 |
|  | 4 | 70.8 [57.9-98.8] | 80.8 [37.5-100] | 0.831 | >1 | 28 |
|  | 5 | 77.4 [27.1-100] | 89.5 [66.9-100] | 0.911 | >1 | 26 |
|  | 6 | 100 [56.4-100] | 100 [61.2-100] | 0.455 | >1 | 25 |
|  | 7 | 100 [66.2-100] | 100 [47.1-100] | 0.195 | >1 | 19 |
|  | **Amplitudes** |  |  |  |  |  |
|  | 1 mA | 34.6 [6.7-75] | 48.1 [23.1-91.7] | 0.042 | 0.231 | 30 |
|  | 2 mA | 80 [56-97.9] | 91.7 [64.4-100] | 0.125 | 0.344 | 27 |
|  | 3 mA | 100 [74.1-100] | 100 [71.3-100] | 0.447 | 0.82 | 21 |

**Supplementary Table S7:** Pairwise Comparisons –Subscores. Only significant differences are shown.

| Outcome | Condition | Median Improvement<br>130 Hz [IQR] | Median Improvement<br>185 Hz [IQR] | p-Value<br>(uncorr.) | p-Value<br>(FDR) | N |
| --- | --- | --- | --- | --- | --- | --- |
| **ET** |  |  |  |  |  |  |
| Postural Tremor<br>[ACC]<br>Improvement<br>[%] | TEED<br>Step 3 | 85.5 [50.5-100] | 62.5 [33.3-97.8] | 0.002 | 0.047 | 28 |
| **PD** |  |  |  |  |  |  |
| Akinetic-rigid<br>Symptoms<br>[Clinical]<br>Improvement<br>[%] | 1mA | 25 [14.3-50] | 50 [37.5-75] | <0.001 | <0.001 * | 30 |
| Akinetic-rigid<br>Symptoms<br>[Clinical]<br>Improvement<br>[%] | 2mA | 66.7 [50-87.5] | 80 [71.4-100] | 0.002 | 0.006 * | 27 |

**Supplementary Table 8:**
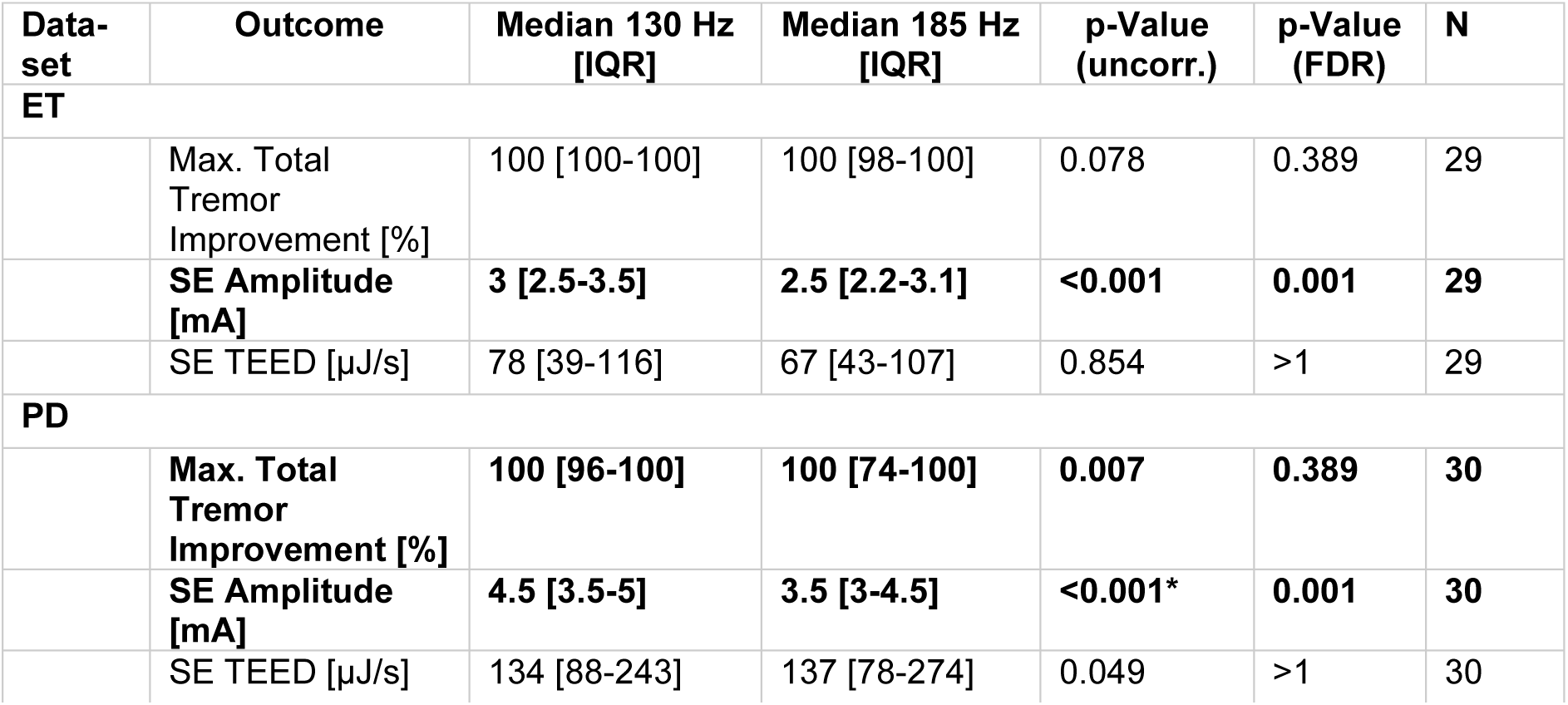
Side Effect Comparisons. Maximal total Tremor Improvement 0.5 mA below the individual side-effect (SE) threshold. Abbreviations: SE = Side-effect, TEED = total electrical energy delivered.

**Supplementary Table 9:** Observed Side Effects for ET and PD with both stimulation frequencies. Abbreviations: ET = Essential Tremor, PD = Parkinson’s Disease.

| Side Effect | ET |  | PD |  |
| --- | --- | --- | --- | --- |
|  | 130 Hz | 185 Hz | 130 Hz | 185 Hz |
| Dysarthria | 17 | 19 | 13 | 12 |
| Ataxia | 10 | 11 | - | - |
| Paresthesia | 5 | 3 | 5 | 10 |
| Dizziness | 3 | 2 | 2 | 2 |
| Troubled Vision | 3 | 2 | - | - |
| Capsule | 1 | 1 | 9 | 10 |
| Unspecific Discomfort | - | - | 2 | 2 |
| Dyskinesia | - | - | 1 | - |

**Supplementary Figure S1:**
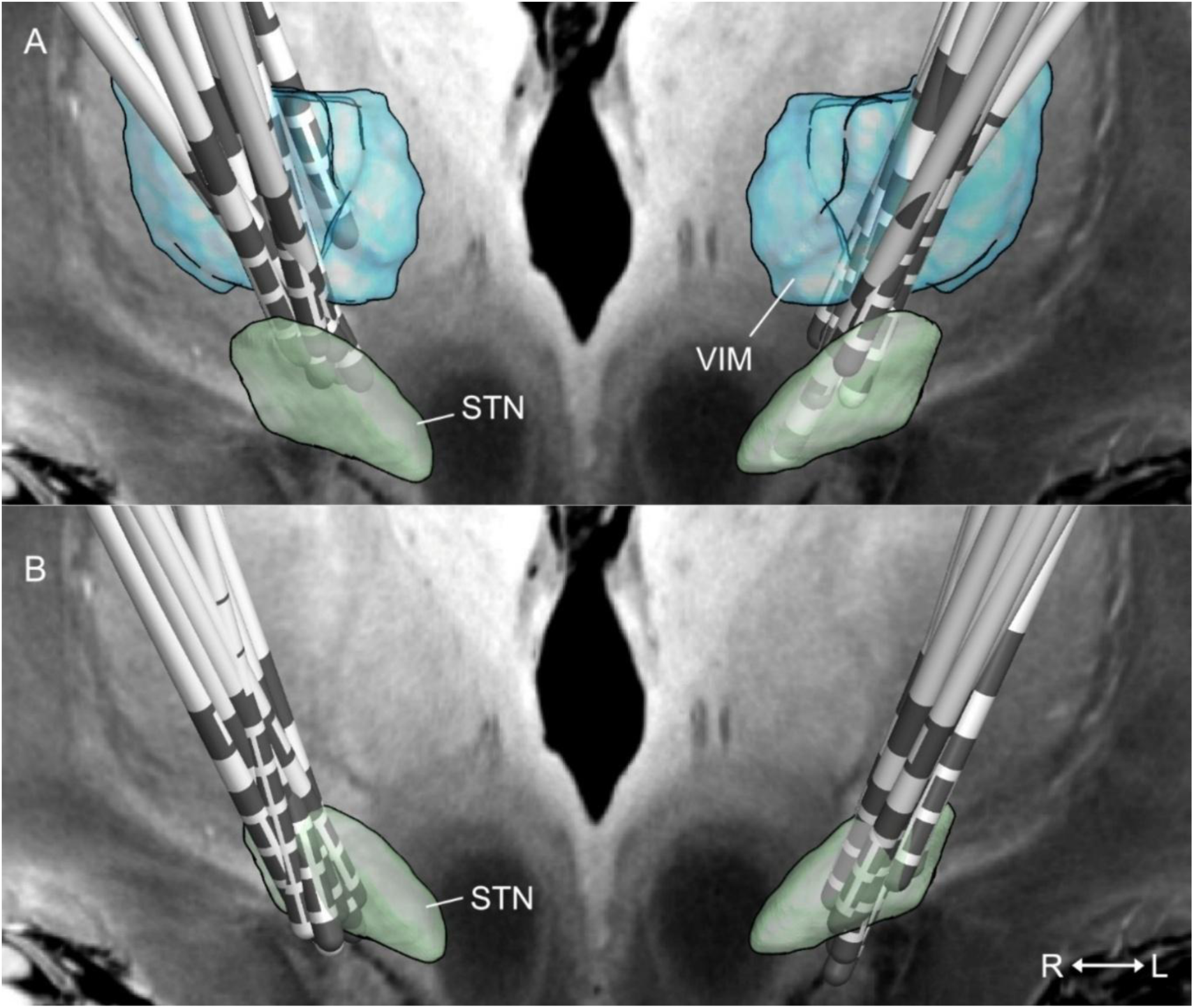
Lead Positions in MNI ICBM 2009b space, illustrated with relevant anatomical structures. Postoperative CT images (IQon Spectral CT, iCT 256, Brilliance 256, Philips Healthcare, Best, The Netherlands) were linearly co-registered to preoperative magnetic resonance imaging (3-Tesla Philips Ingenia® Scanner, Philips, Amsterdam, The Netherlands) using advanced normalization tools (ANTs).^3^ A brain shift correction step was applied as implemented in Lead-DBS. Then, images were non-linearly normalized into standard space using ANTs and the “Effective: low variance, Default” setting. The results of each pre-processing step were visually inspected regarding accuracy and refined if necessary. DBS leads were automatically reconstructed with the PaCER algorithm,^4^ and manually refined. The orientation of the directional leads was determined using DiODE^5^ (A) Essential Tremor Cohort (B) Parkinson’s Disease Cohort Abbreviations: STN = Subthalamic Nucleus, VIM = ventral intermediate nucleus.

